# Meta-Analysis of Risk of Vaccine-Induced Immune Thrombotic Thrombocytopenia Following ChAdOx1-S Recombinant Vaccine

**DOI:** 10.1101/2021.05.04.21256613

**Authors:** Benjamin TB Chan, Pavlos Bobos, Ayodele Odutayo, Menaka Pai

## Abstract

**Context:** Vaccine-induced immune thrombotic thrombocytopenia (VITT) has been reported after administering ChAdOx1-S recombinant COVID-19 vaccine (marketed as Vaxzevira™ by Astra-Zeneca, Covishield™). Estimates of incidence vary between countries, due to different age distributions chosen, case definitions and choice of denominator (persons vaccinated vs immunizations given). This study clarifies these estimates by pooling data from ten countries and examining differences by age group.

**Methods:** We examined case reports, press releases and immunization data and calculated pooled estimates of VITT incidence using random effects models. Sensitivity analyses considered different combinations of countries and varying assumptions on time between vaccination and reporting of cases.

**Results:** Pooling all countries, VITT incidence was 0.73 per 100,000 persons receiving first dose of Covishield/Vaxzevira [95% CI .43,1.23]. Incidence for age 65 and over was 0.11 per 100,000 persons [95% CI .05-.26], and significantly higher among those under age 55: 1.67 per 100,000 persons [95% CI 1.30-2.14] in the UK, 5.06 per 100,000 persons in Norway [95% CI 2.16, 11.86]. The latter had the best data on counts of persons vaccinated. Incidence for age 55 to 64 years was 0.34 [95% CI 0.13, 0.85] in the UK, lower than for under age 55.

**Conclusion:** VITT is a rare vaccine-associated adverse event. Incidence estimates vary between jurisdictions. However, even the highest reported incidence from Norway is low – and in settings with high community transmission, lower than risk of serious outcomes associated with Covid-19. Policymakers and individuals can use these data to calculate risk-benefit ratios and better target vaccine distribution.

**Essentials:**

- This paper measures risk of vaccine-induced immune thrombotic thrombocytopenia (VITT) after ChAdOx1-S recombinant COVID-19 vaccine
- Pooled estimates of incidence were calculated with a random effects model based on data from 10 countries
- Overall risk is 1 in 139,000; for age 65 and over, about 1 in 1,000,000; for age under 55, between 1 in 20,000 to 60,000
- VITT risk is low and varies by age. These data can inform policies around vaccination distribution.

## Introduction

Vaccine-induced immune thrombotic thrombocytopenia (VITT) or vaccine-induced prothrombin immune thrombocytopenia (VIPIT) has been reported following administration of the ChAdOx1 nCov-19 vaccine, a recombinant chimpanzee adenoviral vector encoding the spike glycoprotein of SARS-CoV-2, marketed as Covishield™ (Serum Institute of India PVT Ltd) and Vaxzevira™ (Astra Zeneca). Initial case reports of VITT in Europe were characterized by thrombocytopenia and unusual clots, such as cerebral sinus venous thrombosis and splanchnic thrombosis; soon after, it was realized that these clots can occur in any arterial or venous territory.^1^ Antibodies to platelet factor 4 were later identified in several patients, suggesting a clotting mechanism analogous to heparin-induced thrombocytopenia. Most initially reported patients were under age 55 although some cases were reported in older age groups. Subsequently, numerous countries imposed age restrictions on the vaccine, while others stopped its use altogether^2,3^.

The decision to continue using the vaccine depends on the risk-benefit ratio, which in turn depends on age. The infection mortality ratio of Covid-19 rises dramatically after age 50, while the risk of VITT drops above age 55. There is considerable disagreement between countries regarding the age at which risks outweigh benefits, ranging from age 30 in UK^4^, 50 in Australia^5^, 55 in France^6^ and 60 in Germany^7^, Spain^8^ and the Netherlands^9^. It is important to have a clear estimate of risk of blood clots for different age categories. To date, there has been considerable uncertainty about the true incidence. Earlier estimates were as low as 1 in 2,000,000 persons.^10^ The European Medicines Association reported an incidence of 1 in 290,000 persons and later revised this to 1 in 153,000 persons.^11^ Uncertainty may be due to varying case definitions; different age distributions of vaccine recipients in different countries and changes in these distributions over time; and variations in whether persons vaccinated or vaccines given is used in the denominator.

This paper pools case reports and vaccination data from different countries and time points to estimate the age-specific incidence, so that countries and individuals can make the best risk assessment regarding use. Our hypothesis was that incidence would vary by age group.

## Methods

A meta-analysis was performed to synthesize all the available evidence of the incidence of VITT. We examined data up until April 15, 2021 from countries which had publicly reported VITT-like cases and had estimates of numbers of prior vaccinations. We excluded countries which had not yet approved the vaccine (United States, Switzerland), elected to not use it (New Zealand, Israel), had not publicly reported data within the study period (Belgium, Greece, Poland, Romania, Bulgaria, Ireland), or had some cases but no reported vaccination counts (Finland, Austria). We examined information from regulatory authorities, press releases, scientific journals, and media reports. Information was of three types: vaccination policies describing the age groups and type of individuals targeted for Astra Zeneca vaccination; the number of cases and the age distribution of cases; and number of persons vaccinated.

We included only reports of VITT-like cases with thrombosis associated with thrombocytopenia shortly after vaccination. Cases with only clotting (e.g., deep vein thrombosis, pulmonary embolism or cerebral venous sinus thromboembolism) and no documented thrombocytopenia were excluded. Vaccination with Covishield/Vaxzevira in the countries examined typically began in January to February 2021.

To estimate VITT incidence, we matched the announcement date of cases to the number vaccinations given approximately two weeks prior to the date. This was based on a UK case series that reported median days to admission after vaccination of 12 with right-skewing.^12^ We used number of persons receiving first vaccine dose as the denominator, as this appears to be the moment of greatest risk. (In the UK, which had the most experience with administering second doses, 99% of cases as of April 5 had occurred on the first dose). We categorized countries according to the persons targeted for vaccination during the period of this study: those under 65; those aged 55 to 64; and those offering it to a mix of persons over and under age 65. These policy differences arose from fact that Covishield/Vaxzevira initially had only limited efficacy data from randomized trials on the 65 and over population, which led to some but not all countries restricting its use to those under 65.

Incidence were derived using a logistic transformation and exact confidence intervals were derived using Clopper-Pearson confidence intervals. In the case of one rate reported as zero, a right-side confidence interval was estimated assuming Poisson distribution at value such that the probability of being greater than zero was 95%. Individual incidences were then pooled using an inverse variance random effects meta-analysis of proportions. Results were expressed per 100,000 people. Analyses were conducted using Stata (Version 16) and R (Version 3.6.1).

Next, we estimated age-specific incidence. For the 65 and over population, we used data from the UK where case reports were available in March and most of this population had already been vaccinated by then. For the 55 to 64-year group, we reported separately the incidence for Canada, which de facto had administered this vaccine primarily to this group during this study period^13,14,15,16^, and compared this to data from the UK and Norway. For the under 55 age group, data were available from Norway and the UK.

### Sensitivity analysis and assumptions

Sensitivity analyses were conducted for the age-specific incidence, where we presented, for each age group, a low, medium and high estimate under varying assumptions. In the case of UK data, we examined data using alternate specifications of the “look-back” period (time interval between reporting of cases and time point for counting persons vaccinated), including just shorter and longer than the 12-day median reported in the country.^12^ We also examined case report data from three time points (March 18^17^, 31^18^ and April 4^19^) and tested varying assumptions on how to divide observed cases during these time points into different age categories. In general, we relied on a detailed case series study on the initial 23 patients in the UK with data on age of each case to guide allocation of cases to different age groups.^12^ After examining variations on the above, we identified the highest and lowest estimates observed. Table 2 below lists assumptions for each scenario in more detail.

A similar approach was used to estimate the under-55 and 55- to 64-year incidence in Norway. Information was available on all five cases^20^ as well as Vaxzevira doses given and the age distribution of all vaccines given (including Vaxzevira, Pfizer-BioNTech and Moderna), but not on the age distribution for Vaxzevira doses given specifically^21^. Given that Pfizer-BioNTech and Moderna were targeted to the elderly and Vaxzevira only to those under 65 (mainly young healthcare workers), we assumed that the observed age distribution of vaccines for the under 55 and 55 to 64 year age groups applied to Vaxzevira.

Additional modelling assumptions were made for the UK data. VITT cases were reported for the UK but vaccinations were given for England only.^22^ We assumed that vaccination rates in Scotland, Wales and Northern Ireland were similar and grossed up the English vaccination numbers according to population size. Secondly, age-specific data were available only for all vaccinations given. We estimated the number of Vaxzevira shots given by accounting for information indicating that Vaxzevira for accounted for 22% of vaccines from December 8 to January 24, 59% February 7-28 and 99.5% from March 1 to March 14.^23^ Lastly, we assumed that these proportions applied equally to all age groups; The UK’s National Vaccine plan proposed using either Vaxzevira or Pfizer-BioNTech in its roll-out with no distinction as to which should be used for certain groups.^24^

As an additional component to the sensitivity analysis, we classified countries according to their data quality. Norway and Denmark had high data quality because the vaccination program was put on hold and then cancelled after initial cases were announced^2,3^ hence, the population vaccinated can be measured with greater certainty. Countries with “medium data quality” had weekly vaccination counts, allowing for a two-week look-back period. A country’s data quality was “limited” if the number of persons vaccinated had been announced but no information was available regarding the time point at which this count was measured. We ensured that data from Norway, as a high data quality country, appeared in the sensitivity analysis for age-specific incidence, and for estimates of overall incidence, we conducted a pooled estimate examining only the two high-data-quality countries.

## Results

Table 1 lists, for each country the number of cases, deaths and number of vaccinations given as of some reference date when cases were announced, and the level of data quality.

**Table 1:**
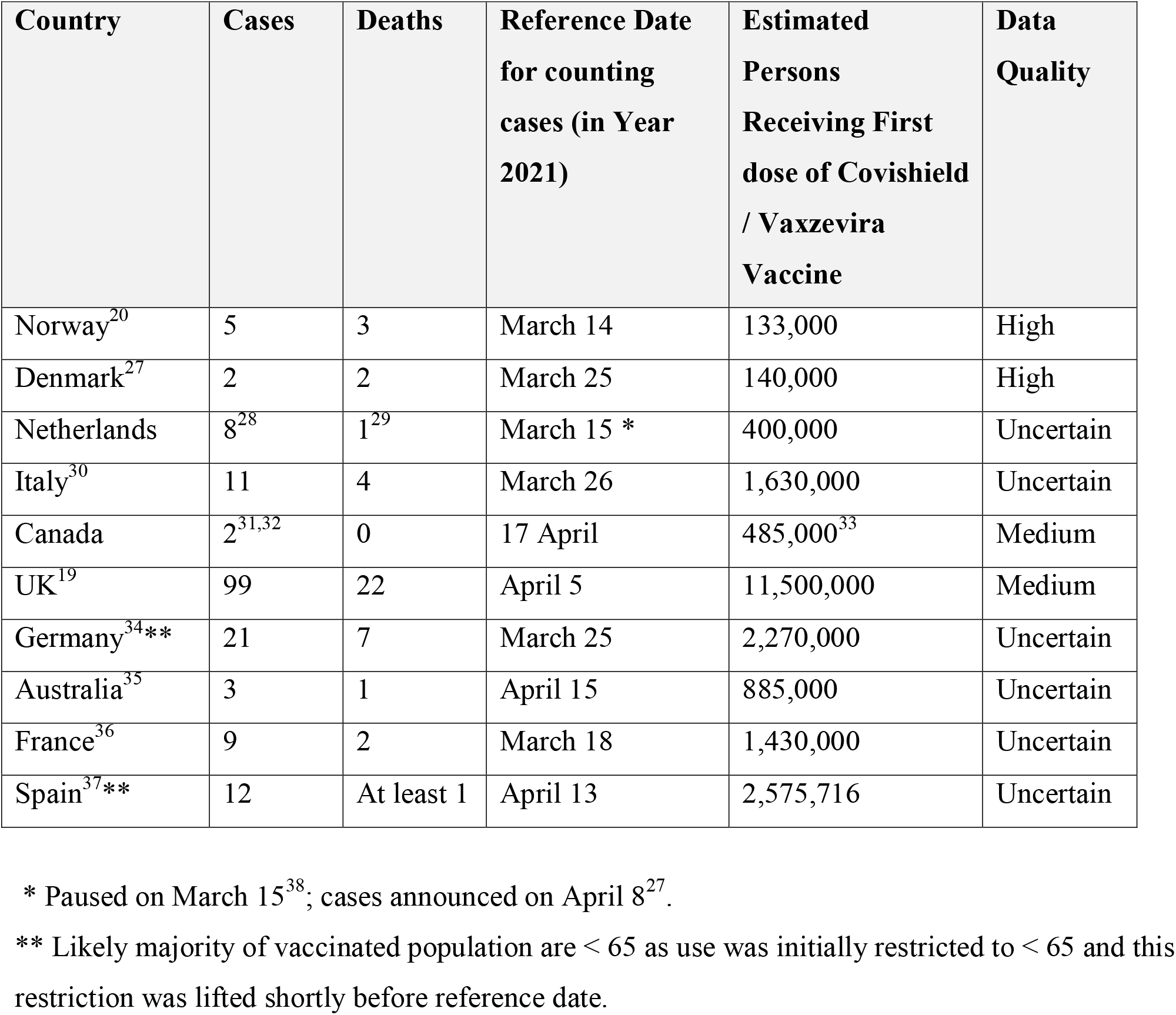
Vaccine-Induced Thrombotic Thrombocytopenia Cases and Deaths by Country.

| **Country** | **Cases** | **Deaths** | **Reference Date for counting cases (in Year 2021)** | **Estimated Persons Receiving First dose of Covishield / Vaxzevira Vaccine** | **Data Quality** |
| --- | --- | --- | --- | --- | --- |
| Norway<sup>20</sup> | 5 | 3 | March 14 | 133,000 | High |
| Denmark<sup>27</sup> | 2 | 2 | March 25 | 140,000 | High |
| Netherlands | 8<sup>28</sup> | 1<sup>29</sup> | March 15 * | 400,000 | Uncertain |
| Italy<sup>30</sup> | 11 | 4 | March 26 | 1,630,000 | Uncertain |
| Canada | 2<sup>31,32</sup> | 0 | 17 April | 485,000<sup>33</sup> | Medium |
| UK<sup>19</sup> | 99 | 22 | April 5 | 11,500,000 | Medium |
| Germany<sup>34**</sup> | 21 | 7 | March 25 | 2,270,000 | Uncertain |
| Australia<sup>35</sup> | 3 | 1 | April 15 | 885,000 | Uncertain |
| France<sup>36</sup> | 9 | 2 | March 18 | 1,430,000 | Uncertain |
| Spain<sup>37**</sup> | 12 | At least 1 | April 13 | 2,575,716 | Uncertain |

\* Paused on March 15<sup>38</sup>; cases announced on April 8<sup>27</sup>.
\*\* Likely majority of vaccinated population are < 65 as use was initially restricted to < 65 and this restriction was lifted shortly before reference date.

Figure 1 contains information from the pooled estimates. The incidence pooled for all countries was 0.73 per 100,000 persons or 1 in 137,000 persons. For the under-65 age group, the pooled estimate for VITT incidence was 1.60 per 100,000 or 1 in 63,000 persons. As a sensitivity analysis for this age group, we calculated a pooled estimate for the two countries with high data quality (Norway and Denmark). This incidence was 2.71 per 100,000 persons (95% confidence interval 1.10 to 6.67), or 1 in 37,000 persons.

**Figure 1:**
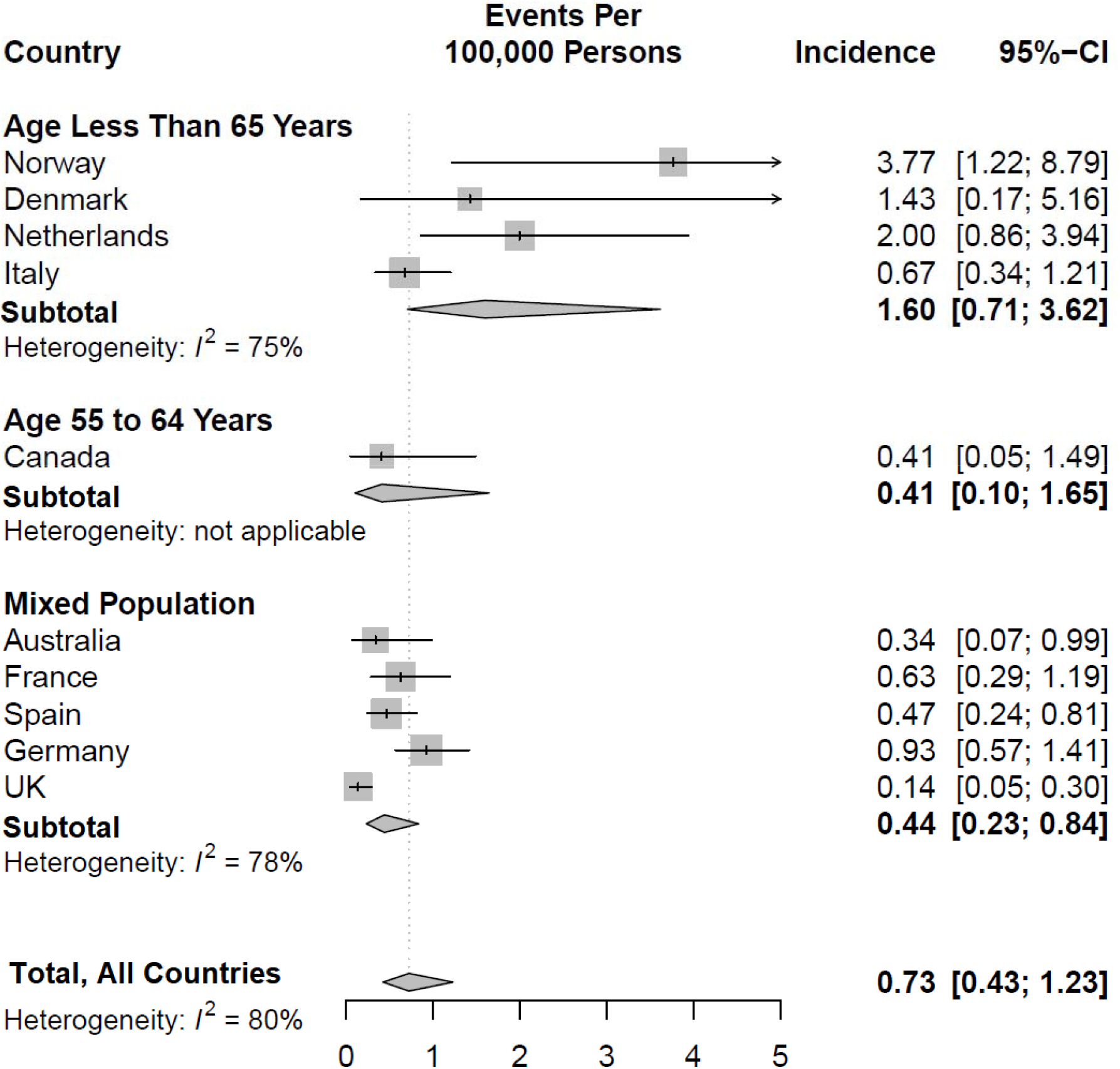
Pooled Estimate of Incidence of Vaccine-Induced Thormbotic Thrombocytopenia using Random Effects Model. Forest plot presenting the pooled incidence of vaccine induced thrombotic thrombocytopenia in countries administering the ChAdOx1-S recombinant vaccine. Each square represents the incidence for a given country. The size of the square is proportional to the weights used in the meta-analysis and the horizontal lines indicating the 95% confidence intervals. The solid diamond within each subgroup indicates the overall pooled incidence and the bottom solid diamond indicates the pooled estimate across all countries. The dashed vertical line corresponds to the overall pooled incidence.

For the mixed age population, the pooled estimate of incidence is 0.44 per 100,000 persons or one in 227,000 persons. As a sensitivity analysis, we excluded the UK which had a disproportionately high percentage of elderly persons in the sample compared to other countries, due to its very successful vaccination campaign in early 2021. This incidence was 0.62 per 100,000 (95% confidence interval 0.41 to 0.94) or 1 in 161,000.

Age-specific incidences are reported in Table 2. There was considerable variation in the medium and high estimates for the 55-year-old age group, ranging from 1 in 60,000 persons in the UK data to 1 in 20,000 persons for Norway, which had high data quality. The incidence was significantly lower in the 55 to 64 age group compared to age 55 in the medium and high scenarios, but confidence intervals were wide. The incidence was lowest in the age 65 and over category, about 1 in a million, and there was relatively little variation between the high and low scenarios. These incidences were significantly different from the incidence for the under-55 age group.

**Table 2:** Incidence of Vaccine-Induced Thrombotic Thrombocytopenia by Age Group with Sensitivity Analysis.

| Scenarios for Sensitivity Analysis | Estimated cases | Estimated population | Cases per 100,000 and 95% confidence interval | Assumptions for Sensitivity Analysis |
| --- | --- | --- | --- | --- |
| Age 55 and under |  |  |  |  |
| UK: low estimate | 61.8 | 5,800,000 | 1.06<br>[0.83,1.36] | 79 cases, March 31; apply 18/23 proportion from case series; 10-day look-back |
| UK: high estimate | 61.8 | 3,700,000 | 1.67<br>[1.30,2.14] | Same as above but 17-day look-back |
| Norway | 5 | 99,000 | 5.06<br>[2.16,11.86] |  |
| Age 55-64 |  |  |  |  |
| Norway | 0 | 34,000 | 0<br>[0,8.8] |  |
| UK: high estimate | 4.16 | 1,230,000 | 0.34<br>[0.13,0.85] | 79 cases, March 31; apply ratio of 1/19 from case series assuming no recent cases < 65; 17-day look-back |
| Canada | 2 | 485,000 | 0.41<br>[0.11,1.50] | Vaccination initially targeted to 60-64 or 55-64 during study period in most provinces. |
| Age 65 and over |  |  |  |  |
| UK: low estimate | 4.0 | 4,650,000 | 0.086<br>[0.033,0.22] | 30 cases reported, Mar 24; assume 4 cases >= 65 (as reported in 23 case series); 10-day look-back |
| UK: medium estimate | 5.22 | 4,400,000 | 0.11<br>[0.05,0.26] | Apply proportion of 4/23 found in case series to 30 cases; 17-day look-back |
| UK: high estimate | 5.22 | 4,400,000 | 0.12<br>[0.05,0.27] | Apply proportion of 4/23 found in case series to 30 cases; 24-day look-back |

“Look-back” is time interval between date when cases officially were reported and reference date used to count number of people vaccinated; longer look-backs used in high estimates. “Case series” refers to published findings on age distribution of initial 23 cases found in UK.<sup>12</sup>

### Interpretation

The overall VITT incidence of 1 in 137,000 persons across all countries is consistent with the more recent estimates from the European Medicines Agency noted above. There were wide variations between countries in their estimates of incidence, likely due to differences in reporting methods and the age distribution of persons vaccinated. Hence, the reporting of an “overall incidence” level is fraught with pitfalls. It is recommended that incidence by major age groups be quoted going forward, as these are more transparent.

There were important differences by age group. Based on UK data, the thrombosis incidence over age 65 is very low, approximately one in a million. This may explain why VITT-like cases were not detected sooner in the UK, which had implemented an aggressive vaccination strategy initially targeting the elderly and those in care homes. Cases increased steadily from March onwards as younger cohorts began to be vaccinated. The very low incidence observed in the elderly suggests that this is a reasonable age group to target for receiving Covishield/Vaxzevira vaccine. However, one must consider the possibility that this rate was under-estimated because the elderly were vaccinated first in the UK and cases might not have been missed given that VITT had not yet been recognized. Estimates needs to be confirmed in countries using the vaccine in the elderly in the near future.

The incidence of VITT is considerably higher in the under 55 age group, with medium and high estimates ranging from 1 in 60,000 persons to 1 in 20,000 persons. Policymakers should consider the latter as a worst-case scenario in their policies on which patients should receive the vaccine, given that this estimate was based on Norway which had among the best data quality. It should be noted, however, that even in this worst-case scenario, the incidence is less than or comparable to that for other common, routine activities such as driving (for example, 1 in 3,000 risk of serious injury requiring hospitalization, 1 in 19,000 risk of death per year in Canada^25^). To put these risks further into perspective, the risk of venous thromboembolism is high among hospitalized Covid-19 patients,^26^ 21% if non-ICU and 31% if in ICU. A pre-print publication further suggests that the risk of CSVT and portal vein thrombosis following Covid-19 is 35.3 and 393.3 per million persons respectively; these rates together represent a risk that is 9 times higher than the highest estimate of VITT incidence identified in this study.^27^

VITT incidence in the 55 to 64 age group was 1 in 300,000 persons in the medium scenario, and significantly lower than in the under age 55 group. However, confidence intervals were wide. Getting more precision for this incidence will be important as more data become available, given that several countries have restricted vaccinations to the over 55 or 60 age groups. The desirability of these policies depends on this estimate.

Policymakers can use this data to determine which patients should be eligible to receive the vaccine in their jurisdiction. This decision should also consider the prevalence of Covid-19 in the community, an individual’s risk of being exposed (due, for example, to being an essential worker), and age, gender, presence of underlying health conditions which may increase the infection fatality rate. Groups identified as having a higher risk of death from Covid-19 compared to VITT should receive the vaccine. The availability of alternative vaccines should also be considered; if there is a delay in the supply of alternative vaccines while cases are being transmitted rapidly in the community, then the risk of waiting for alternatives needs to be weighed against the VITT risks identified here.

### Limitations

We believe the most important limitation of this analysis is the potential for undercounting of cases, particularly in the early stages of vaccination campaigns before VITT was recognized as a complication. The results in the UK in the over-65 group should be interpreted with caution since this was one of the first groups vaccinated worldwide with this vaccine. While we calculated a low assumption for each age-specific incidence, it may be prudent for policymakers to use the medium and high assumptions for planning purposes.

VITT incidence could also be underestimated due to improper timing of when cases and vaccines are counted leading to errors. As noted, given the typical 12-day delay between vaccination and hospital admission, vaccinations should be counted about two weeks prior to the case date. Ideally, a cohort of vaccinated patients should be described and then all cases coming forward from this group should be counted. In practice, this was possible only for Norway and Denmark as noted above.

Except for three published case reports, data were limited on the exact age distribution of cases. We were not able to obtain access to individual-level data in adverse events databases. Readily accessible data on age distribution of vaccines given, the type of vaccine given and breakdowns by week were available only in certain jurisdictions such as the UK, Norway and Canada, and even in these instances some assumptions had to be made regarding missing information.

Because this study was conducted early after the recognition of VITT as a complication of vaccination, case counts were relatively low and hence, confidence intervals were wide. This study should be replicated in the near future to obtain more precise estimates, and ideally, calculations should be updated regularly and posted publicly.

## Conclusion

This study confirms that VITT after the ChAdOx1 nCov-19 vaccine is particularly rare in those over age 65 (1 in 1 million), but more common under age 55 (from 1 in 20,000 to 60,000 persons). This information can be used by policymakers in risk-benefit calculations, comparing risks of vaccine associated adverse events to benefits of avoiding COVID-19 infection, and in targeting vaccination campaigns accordingly. It can also help the general population frame the magnitude of the risk of VITT, relative to other common baseline risks in their environment.

## Data Availability

All data sources used in this analysis are listed in references.

## Addendum

### Authorship Details

B. Chan conceived of study topic, designed methods, obtained data and was lead author of manuscript. P. Bobos performed the data synthesis and meta-analysis, approved of final version for submission. A. Odutayo performed data synthesis and meta-analysis, reviewed manuscript for important context, approved of final version. M. Pai made contributions to the interpretation of data for the work; assisted with revisions; approved of final version for submission.

## Acknowledgements

Thanks Dr. Michael Schull for his comments in an earlier draft of this paper, and to Susmitha Rallabandi for administrative support. Responsibility for accuracy and content rests with the authors.

